# Modified Ghost System combining action observation and vibration stimulation for recovery after distal radius fracture surgery: A single-arm clinical feasibility study protocol

**DOI:** 10.64898/2026.07.16.26358289

**Authors:** Akito Kano, Yosuke Akiyama, Yoshi-Ichiro Kamijo, Toyohiro Hamaguchi

## Abstract

Distal radius fractures (DRFs) can delay return to activities of daily living and social participation because of postoperative pain, temporary joint immobilization, and limited wrist and forearm range of motion. The Ghost System developed at Saitama Prefectural University, Japan, combines visual action observation with tendon vibration stimulation and has shown potential as an adjunct to conventional rehabilitation. This Study Protocol describes a modified Ghost system intended to improve clinical implementation by replacing the head-mounted virtual reality display with iPad-based action observation and by using a wristband-type vibrator. This single-center, single-arm, open-label feasibility trial will enroll 10 adults after palmar locking plate fixation for DRF. The intervention will be delivered twice weekly during outpatient rehabilitation follow-up sessions from the early postoperative period (postoperative days 2-10 after enrollment) through the approved early postoperative rehabilitation period (generally up to postoperative week 8), in parallel with standard rehabilitation practices. Primary feasibility and preliminary clinical outcomes include device fit and acceptability, pain assessed using a 100-mm Visual Analog Scale, and wrist/forearm range of motion. Secondary implementation and safety outcomes include Disabilities of the Arm, Shoulder and Hand (DASH), Patient-Rated Wrist Evaluation (PRWE), Hand20 Questionnaire (HANDS-20), EuroQol 5 Dimensions 5 Levels (EQ-5D-5L), body ownership and hand-illusion questionnaires, setup time, setup errors, adherence, adverse events, and device incidents. We hypothesize that the modified Ghost system will be feasible and acceptable for early postoperative outpatient rehabilitation and will be delivered without serious device-related adverse events. Clinical outcomes will be summarized descriptively to inform a future controlled study rather than to establish efficacy.

## Introduction

A distal radius fracture (DRF) is a common fall-related injury in which external force is applied to the distal radius; DRFs account for a significant proportion of adult fractures [1]. After palmar locking plate fixation, early rehabilitation may begin during the postoperative period, but pain, temporary immobilization, edema, and limited wrist and forearm range of motion (ROM) can delay hand use and recovery of activities of daily living (ADLs) [2,3,4].

Pain after DRF surgery may also promote fear avoidance, catastrophic thinking, anxiety, and disuse, which can slow functional recovery and increase disability [5,6]. Patients with DRF may underestimate their own ROM recovery, and lower subjective recovery has been associated with poorer grip strength and hand function [2]. Strategies that reduce pain and promote safe early sensorimotor engagement may therefore be useful adjuncts to standard rehabilitation.

Saitama Prefectural University, Japan, developed the original Ghost System (Patent No. 6425355), which combined vibrotactile stimulation with visual stimulation using a head-mounted virtual reality display and a vibration motor [8,14]. Previous studies of action observation, motor imagery, and tendon vibration-induced kinesthetic illusion suggested that visual and proprioceptive inputs can engage sensorimotor processes while limiting the need for strenuous voluntary movement [7,9–13]. The present Study Protocol does not directly evaluate cortical motor excitability or motor representation. Therefore, the modified Ghost System is positioned as a sensorimotor intervention that combines iPad-based visual action observation and vibration-induced proprioceptive input to support kinesthetic illusion, body ownership-related sensations, sense of agency, motor intention, and mild voluntary movement. Previous studies suggested that adding the Ghost System to conventional rehabilitation programs may improve pain, ROM, and function in patients with DRF, but the original device has implementation barriers in routine outpatient care, including head-mounted display burden, portability, setup time, and cost [14].

This Study Protocol describes a modified Ghost System developed to reduce these implementation barriers. The modified system uses iPad-based action observation and a wristband-type vibrator placed over the extensor carpi ulnaris (ECU) tendon on the dorso-ulnar aspect of the distal forearm/wrist on the unaffected side. This intervention delivers synchronized visual and vibration stimuli while promoting motor intention and mild, comfortable, screen-synchronized voluntary movement. Body ownership-related sensations, kinesthetic illusion, and sense of agency are assessed exploratorily using patient-reported questionnaires.

The aim of this Study Protocol is to evaluate the feasibility, safety, and acceptability of adding the modified Ghost System to standard outpatient rehabilitation follow-up after DRF surgery and to describe preliminary changes in pain, wrist/forearm ROM, and patient-reported outcomes. We hypothesize that the intervention will be feasible and acceptable, delivered without serious device-related adverse events, and show favorable preliminary changes in pain and ROM. Efficacy and causal effects cannot be established because this is a single-arm feasibility study with 10 participants.

## Materials and Methods

The full step-by-step intervention protocol is available online at protocols.io (https://dx.doi.org/10.17504/protocols.io.5qpvoem2bl4o/v1). The subsections below summarize the study design, population, outcomes, device components, intervention procedure, safety monitoring, data management, and statistical analysis.

### Study design, clinical setting, and registration

This is a single-center, single-arm, open-label, non-randomized clinical feasibility trial protocol. No concurrent control group or formal historical-control comparison will be used for efficacy testing. All participants will undergo standard outpatient rehabilitation follow-up as prescribed by the attending clinician, and the modified Ghost System will be added as an adjunct intervention. The trial is registered in the University Hospital Medical Information Network Clinical Trials Registry (UMIN-CTR; UMIN000061163). The ethics-approved study protocol is version 1.1 dated March 20, 2026.

This study is being conducted in the outpatient rehabilitation setting of Dokkyo Medical University Saitama Medical Center, Saitama, Japan. Before enrollment, a study researcher explains the ethics committee-approved participant information sheet, confirms the participant’s understanding, and obtains written informed consent before any study-specific intervention. No genetic or genomic information is collected (Fig 1).

**Figure 1.**
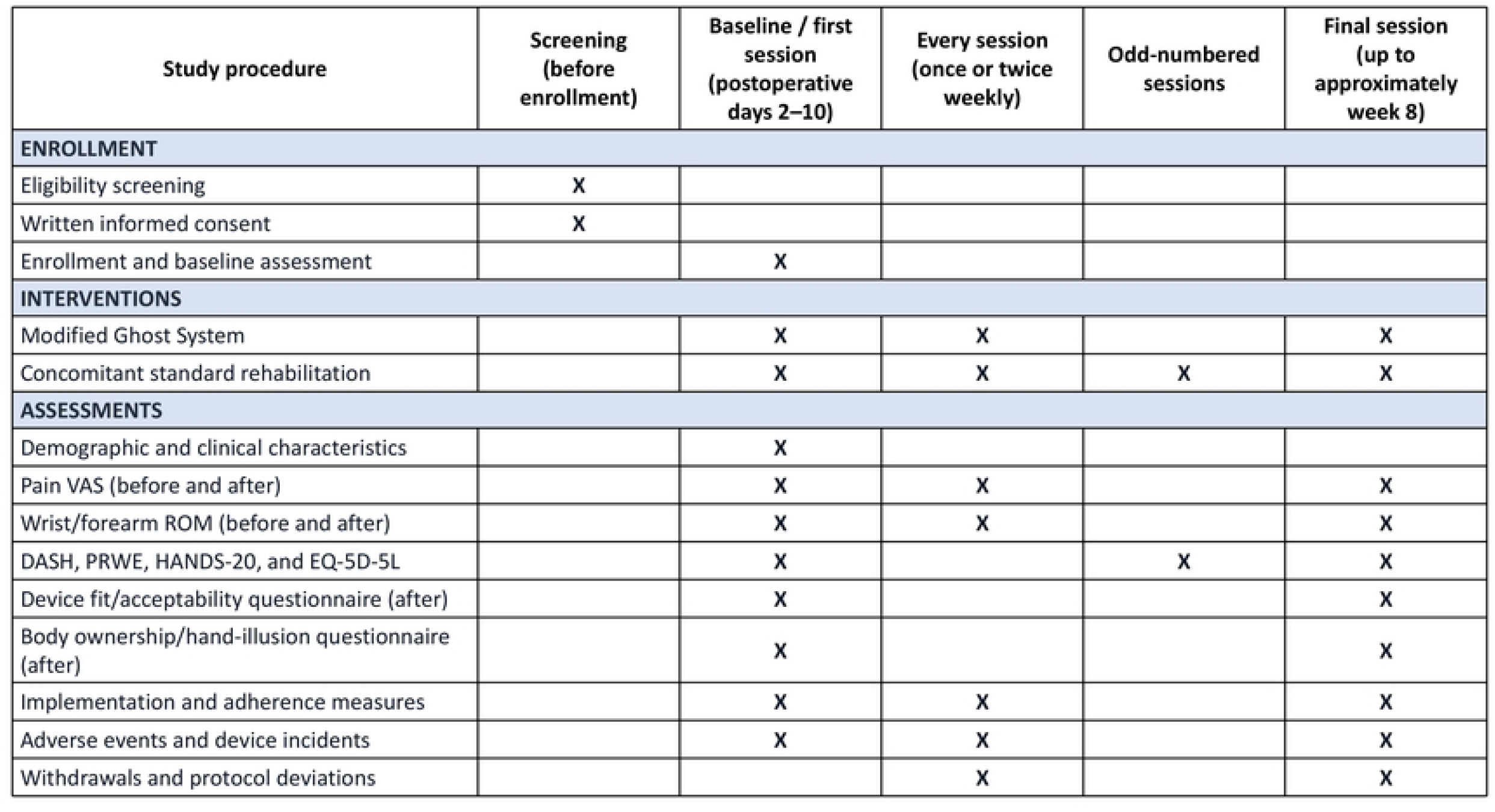
completed SPIRIT schedule. Schedule of enrollment, interventions, and assessments. X, scheduled procedure or 135 assessment. Each session includes the first and final sessions. VAS and ROM are assessed 136 immediately before and after each modified Ghost System session. Patient-reported outcomes are 137 assessed at baseline, before odd-numbered sessions, and at the final session.

This is an investigator-initiated study with no commercial sponsor. Dokkyo Medical University Saitama Medical Center is the coordinating institution. Yoshi-Ichiro Kamijo is the principal investigator responsible for study conduct and supervision. Akito Kano is the study coordinator and study contact and coordinates recruitment, intervention delivery, and data collection.

Patients and members of the public were not involved in the design, conduct, or reporting plans for this study. Participant feedback on device fit and acceptability is collected as a study outcome and is not considered patient and public involvement.

### Participants

The eligible participants are adults aged 18 years or older who have undergone palmar locking plate fixation for DRF, are prescribed outpatient rehabilitation, are willing to use the modified Ghost System, and have provided written informed consent. The exclusion criteria are joint contracture due to a prolonged interval between injury and surgery, suspected moderate cognitive impairment as defined in the approved protocol, more than 10 days since DRF surgery at enrollment, a life-threatening medical condition, or any other reason judged by the investigator to make participation inappropriate.

To assess clinical feasibility, this pilot Study Protocol will enroll 10 patients. No formal sample-size calculation based on efficacy testing was performed because the purpose was to evaluate feasibility, safety, acceptability, and preliminary outcome estimates. A target of 10 participants was selected as a pragmatic sample size for an early single-center feasibility Study Protocol within the approved study period and clinical setting.

Potentially eligible patients attending the orthopedic outpatient service who have been prescribed rehabilitation are screened by the research team against the eligibility criteria. Recruitment will continue until 10 participants have been enrolled or October 31, 2027, whichever occurs first. The intervention is delivered by licensed rehabilitation professionals who have been trained in the study protocol and operation of the modified Ghost System.

### Intervention: modified Ghost System

The modified Ghost System (Device Approval Number: Saitama 202509-2) consists of a wristband-type vibrator and an iPad-based action observation device. The wristband-type vibrator uses a DC 5 V USB-rechargeable vibration motor with an adjustable speed of 3000-4500 rpm, corresponding to approximately 50-75 Hz. The selected speed setting will be recorded for each session. The motor is mounted in a Reebok Sports Wristband Short or equivalent wristband, approximately 8 cm x 8 cm x 1.5 cm.

The visual stimulus is displayed on an iPad placed on a towel-covered rack or equivalent stand on the table. Four pre-prepared videos are to be used: crossed palms with the left thumb on top, crossed palms with the right thumb on top, wrist extension/flexion displayed toward the left, and wrist extension/flexion displayed toward the right. Each movement cycle consists of 5 seconds to the maximum wrist extension or flexion angle and 1 second to return to neutral. A 36-second stimulation block therefore contains six 6-second cycles.

### Intervention procedure

The intervention begins after enrollment and within postoperative days 2-10 and is continued twice weekly during outpatient rehabilitation follow-up through the approved early postoperative rehabilitation period (up to approximately postoperative week 8). Each session is delivered in parallel with standard rehabilitation practices.

The participant is seated on a chair with a backrest in front of a table equipped with the modified Ghost System, with the seat height set to approximately 40 cm. The table and chair are adjusted so that the participant can look down toward the iPad without neck strain. The distance between the eyes of the participant and the iPad screen is set to approximately 15 cm unless this causes discomfort; any deviation is recorded.

A wristband-type vibrator is placed over the ECU tendon on the dorso-ulnar aspect of the distal forearm/wrist on the unaffected side, and the wristband pressure is standardized to 1 N or less [15]. Vibration will be delivered within the approved adjustable range of 3000-4500 rpm, corresponding to approximately 50-75 Hz, and the selected setting will be recorded for each session.

During stimulation, the selected video and vibration are delivered simultaneously. Participants will be instructed to keep the body relaxed, attend to the hand movement displayed on the screen, and gently move the designated hand in synchrony with the screen movement within a comfortable ROM that does not increase pain. They will also be encouraged to perceive the displayed hand as if it were their own hand. The therapist will provide standardized verbal prompts, avoid encouraging forceful or painful movement, and monitor symptoms continuously.

One rehabilitation exercise set consists of three 36-second simultaneous visual-vibratory stimulation blocks separated by 10-second rest periods. Three rehabilitation exercise sets are performed per session, with 60-second rests between sets, unless the participant stops early or the therapist decides that continuation of the rehabilitation exercise would be inappropriate. The total active stimulation time is 324 seconds per session, and the approximate exercise session duration (including within-set and between-set rests) is 504 seconds.

### Concomitant standard rehabilitation

The modified Ghost System intervention is delivered in parallel with standard rehabilitation practices prescribed by the attending clinician. Standard rehabilitation practices may include physical modalities such as heat therapy, whirlpool, and hand incubator for swelling or edema; active and passive ROM exercises for the wrist and forearm; ADL training involving hand use; and home exercise instruction including wrist movement, forearm rotation, tendon gliding, light strengthening, and loading exercises when clinically appropriate. The type and duration of standard rehabilitation practices provided at each visit will be recorded.

### Outcomes

Outcomes are organized into primary feasibility and preliminary clinical outcomes, secondary patient-reported outcomes, implementation and safety outcomes. The primary outcomes are: (1) device fit and acceptability, assessed with an 8-item Numeric Rating Scale questionnaire after the first and final sessions; (2) pain, assessed using a 100-mm Visual Analog Scale (VAS) before and after each modified Ghost System session; and (3) wrist and forearm ROM measured with a goniometer before and after each session. ROM measurements include wrist palmar flexion, dorsiflexion/extension, radial deviation, ulnar deviation, forearm pronation, and forearm supination.

Secondary patient-reported outcomes include the Disabilities of the Arm, Shoulder and Hand questionnaire (DASH), Patient-Rated Wrist Evaluation (PRWE), Hand20 Questionnaire (HANDS-20), EuroQol 5 Dimensions 5 Levels (EQ-5D-5L), and a body ownership/hand-illusion questionnaire adapted from a previous report on embodiment and body ownership [16]. These questionnaires will be collected according to the approved assessment schedule, including baseline, odd-numbered sessions, and/or final assessment as applicable.

Implementation outcomes include adherence, number of completed sessions and rehabilitation exercise sets, setup time from the second session onward, setup errors, corrective instructions, selected video, vibration setting, and protocol deviations. Safety outcomes include adverse events and device incidents from the start to the end of intervention, including pain worsening, fracture-related worsening, skin problems, dizziness, nausea, headache, fatigue, motion sickness, device malfunction, video malfunction, and iPad/rack instability.

### Safety monitoring and discontinuation criteria

A trained rehabilitation professional will remain in the room throughout the intervention. The intervention will be stopped immediately if the participant requests discontinuation; pain markedly worsens; dizziness, nausea, headache, fatigue, or motion sickness occurs; skin irritation or pressure injury is observed; the device malfunctions; or the therapist decides that continued exercise would be inappropriate. After stopping the exercise, vibration will be turned off, the wristband will be removed, the participant will be assessed, standard clinical care will be provided as needed, and the event will be recorded in the adverse event/device incident log.

If study-related health harm occurs, the participant will receive appropriate standard medical care under the usual health insurance system. No special compensation scheme is provided for participation-related harm.

### Data management

All study data will be recorded using study participant codes. Names and other directly identifiable information will not be included in analysis files. Session-level data will include the date, postoperative day/week, standard rehabilitation exercises provided, modified Ghost System completion, number of completed exercise sets, selected video, vibration setting, setup time, setup errors, adverse events, device incidents, and protocol deviations. Missing data and reasons for their absence will be recorded. Data will be stored according to institutional policies and ethics approval, and access will be limited to authorized research personnel.

Study visits are coordinated with scheduled outpatient rehabilitation whenever possible. Attendance, missed sessions, discontinuation, withdrawal, and reasons for missing data are recorded. Data collected before discontinuation or withdrawal will be retained and analyzed when permitted by the participant’s consent and the ethics approval.

### Study monitoring and protocol amendments

The principal investigator oversees study conduct, protocol adherence, data completeness, and safety. A trained rehabilitation professional monitors the participant during every intervention session and reports adverse events and device incidents to the principal investigator. Because this is a small, single-center feasibility trial with no formal efficacy testing or interim analysis, no independent data monitoring committee, steering committee, or endpoint adjudication committee was established. The study undergoes annual continuation review by the ethics committee. Important protocol amendments will be submitted to the ethics committee and, after approval, reflected in the trial registry, study documents, and reports as applicable.

## Statistical analysis

All analyses will be exploratory and descriptive. The feasibility and safety analysis set will include all participants who received at least one modified Ghost System intervention.

Continuous variables will be summarized using mean and standard deviation or median and interquartile range values, depending on distribution of the data. Categorical variables will be summarized using counts and percentages.

Device fit/acceptability, fatigue, discomfort, perceived usefulness, perceived necessity, willingness/ability to continue, acceptability, setup time, setup errors, adherence, withdrawals, adverse events, and device incidents will be summarized descriptively. Pain VAS and wrist/forearm ROM will be summarized at each time point, and within-session pre-post changes and changes from baseline to final assessment will be calculated. Mean or median changes with 95% confidence intervals will be reported where appropriate.

DASH, PRWE, Hand20 Questionnaire (HANDS-20), EuroQol 5 Dimensions 5 Levels (EQ-5D-5L), and body ownership/hand-illusion scores will be summarized at baseline and final assessment, and exploratory changes will be described. Inferential analyses, if performed, will be considered supplementary because the target sample size is 10. If performed, paired t-tests or Wilcoxon signed-rank tests will be used according to data distributional assumptions, but no efficacy conclusions will be drawn. No adjustment for multiple comparisons is planned. Missing data will not be imputed; analyses will be based on available data, and reasons for their absence will be summarized.

### Study status and timeline

Participant recruitment commenced on April 10, 2026. At the time of resubmission in July 2026, participant recruitment and data collection were ongoing; neither stage had been completed, and no interim or final analyses had been performed. Recruitment is expected to be completed by October 31, 2027, or earlier when the target sample of 10 participants has been enrolled. Data collection, including the final participant’s follow-up, is expected to be completed by December 31, 2027. Data analysis is expected to be completed and the results to be available by March 2028. Findings will be disseminated through a master’s thesis, presentation at a relevant scientific conference, and publication in a peer-reviewed journal, and the trial registry will be updated as appropriate.

### Expected results

The modified Ghost System is expected to be feasible and acceptable for early postoperative outpatient rehabilitation follow-up after DRF surgery, with no serious device-related adverse events. This Study Protocol is expected to provide descriptive estimates of adherence, setup time, setup errors, device fit, acceptability, adverse events, pain, wrist and forearm ROM, and patient-reported outcomes. Any observed changes in pain, ROM, DASH, PRWE, Hand20 Questionnaire (HANDS-20), EuroQol 5 Dimensions 5 Levels (EQ-5D-5L), and body ownership/hand-illusion scores will be interpreted as preliminary signals that will be used to inform the design and sample-size planning of a future controlled study, rather than as evidence of efficacy.

## Discussion

This Study Protocol describes a pragmatic, clinically implementable modification of the Ghost System for early postoperative rehabilitation follow-up after DRF surgery. The modified system uses an iPad and a wristband-type vibrator, which may reduce barriers associated with head-mounted virtual reality equipment, improve portability, and simplify implementation in outpatient rehabilitation settings. The current Study Protocol also specifies standardized device placement, stimulus timing, participant instruction, safety monitoring, and session-level implementation measures.

The present Study Protocol has several limitations. First, it is a single-center, single-arm, open-label feasibility study with a target sample size of 10 participants; therefore, it cannot establish efficacy or causality. Second, the natural recovery that occurs after DRF surgery cannot be separated from the effect of the modified Ghost System. Third, the intervention is delivered in parallel with standard rehabilitation practices, so the independent effect of the modified Ghost System cannot be isolated. Fourth, the twice-weekly intervention schedule may be affected by outpatient attendance and participant circumstances; missed or rescheduled sessions will therefore be recorded. Fifth, participant-reported body ownership, illusion, discomfort, and acceptability measures are exploratory and may be influenced by expectation and therapist interaction. These limitations will be considered when interpreting the preliminary clinical outcomes and for planning future controlled studies.

## Data Availability

For Study Protocols: No datasets were generated or analysed during the current study. All relevant data from this study will be made available upon study completion.

## Authors’ contributions

Conceptualization: AK, TH. Methodology: AK, YA, YK, TH. Investigation: AK, YA. Resources: YK, TH. Writing - original draft: AK. Writing - review and editing: YA, YK, TH. Supervision: TH. Project administration: TH.

## Acknowledgements

The authors thank Associate Professor Yuji Koike of the Graduate School of Saitama Prefectural University (Saitama, Japan) for guidance during preparation of this protocol. The authors also thank the staff of the Department of Orthopedics, the Department of Rehabilitation, and the Occupational Therapy Division at Dokkyo Medical University Saitama Medical Center for their cooperation in this study.

## Supporting information

S1 Protocol. Ethics-approved study protocol in the original Japanese (version 1.1, March 20, 2026).

S2 Protocol. English translation of the ethics-approved study protocol.

S3 Checklist. Completed SPIRIT 2025 checklist.

## Ethics declarations

This study is a single-arm, open-label intervention trial registered in the UMIN Clinical Trials Registry (UMIN-CTR; UMIN000061163) and approved by the Ethics Committee of Dokkyo Medical University Saitama Medical Center (Saitama, Japan; Ethical Approval Number: 25123). The ethics-approved study protocol is version 1.1 dated March 20, 2026. Use of the modified Ghost System was approved under Device Approval Number Saitama 202509-2. A study researcher obtains written informed consent from all participants before enrollment. The study is conducted in accordance with the Declaration of Helsinki and applicable institutional regulations. The assessment schedule is shown in Fig 1. No genetic or genomic information is collected.

## Associated content

Detailed step-by-step intervention protocol: https://dx.doi.org/10.17504/protocols.io.5qpvoem2bl4o/v1

## Clinical trial registration

UMIN Clinical Trials Registry Number: UMIN000061163.

## References

1. Nakayama J, Tano K. Treatment results of distal radius fractures using DTSaM. J Insur Med Sci. 2022.

2. Usuki K, Suzuki T, Ueda H, Ishioka T, Hamaguchi T. Underestimated active joint motions in patients with distal radius fractures: An observational study. Journal of Ergonomic Technology. 2021;21(1):29–39.

3. Abe Y, Isago K, Ito T, Shiiki E, Morita H. Functional recovery after distal radius fracture surgery. Orthop Surg Traumatol. 1998;47(3):825–828.

4. Hasegawa M, Yamashita M, Ushi-no-hama M, Nakamichi M, Tajima Y. Relationship between functional impairment and ADL disability after distal radius fractures. Proc Kyushu Phys Ther Occup Ther Conf. 2004.

5. Meulders A. From fear of movement-related pain and avoidance to chronic pain disability: A state-of-the-art review. Curr Opin Behav Sci. 2019;26:130–136. 10.1016/j.cobeha.2018.12.007

6. Hamagami Y, Nakano J, Sakamoto J, Okita M. Mechanisms of inactivity-induced pain and effects of vibratory stimulation in a rat model. Jpn J Basic Phys Ther. 2017;20(2).

7. Imai R, Osumi M, Morioka S. Influence of illusory kinesthesia by vibratory tendon stimulation on acute pain after surgery for distal radius fractures: A quasi-randomized controlled study. Clin Rehabil. 2016;30(6):594–603. 10.1177/0269215515593610

8. Hamaguchi T, Nakamura-Thomas H. Mechanism of the developed sensorimotor therapy device: Synchronous inputs of visual stimuli and vibration to improve recovery of distal radius fractures. Biomed J Sci Tech Res. 2021;38(3):30612–30616. 10.26717/BJSTR.2021.38.006120

9. Kaneko F, Blanchard C, Lebar N, Nazarian B, Kavounoudias A, Romaiguere P. Brain regions associated with a kinesthetic illusion evoked by watching a video of one’s own moving hand. PLoS One. 2015;10(8):e0131970. 10.1371/journal.pone.0131970

10. Abe H, Kaneko F, Shibata E, Kimura T. Effects of visual stimulation using video on kinesthetic illusion and voluntary movement. Jpn J Basic Phys Ther. 2015;18(2).

11. Kawamura T, Shigemasu H. Effects of visual information and depth position on proprioception. TVRSJ. 2016;21(1):141–147.

12. Makino N, Hattori T, Kurachi T, Takasawa M, Iwasa M, Jo Y, et al. Analgesic effects of motor imagery: Comparison of kinesthetic and visual imagery. 2014. 10.14900/cjpt.2014.1330

13. Usuki K, Ueda H, Hamaguchi T, Yamaguchi T, Suzuki T. Action observation intervention using three-dimensional movies improves hand usability in women with distal radius fractures: A nonrandomized controlled trial. PLoS One. 2024;19:e0294301. 10.1371/journal.pone.0294301

14. Narita D, Hamaguchi T, Nakamura-Thomas H. Development and trial of a prototype device for sensorimotor therapy in patients with distal radius fractures. Appl Sci. 2022;12(4):1967. 10.3390/app12041967

15. Honda M, Karakawa H, Akahori K, Miyaoka T, Ooka M. Effects of different vibration conditions on kinesthetic illusion and perception. TVRSJ. 2014;19(4):457–466.

16. Longo MR, Schüür F, Kammers MPM, Tsakiris M, Haggard P. What is embodiment? A psychometric approach. Cognition. 2008;107(3):978–998. doi:10.1016/j.cognition.2007.12.004.

